# Use of the test-negative design to estimate the protective effect of a scalar immune measure: A simulation analysis

**DOI:** 10.1101/2024.11.22.24317757

**Authors:** Ziyuan Zhang, Christopher Brian Boyer, Marc Lipsitch

## Abstract

**Background:** The relationship between antibody levels (more generally, a scalar measure of immune protection) at the time of exposure to infection (so-called exposure-proximal correlates of protection) and the risk of infection given exposure is of central interest in evaluating the evolution of immune protection conferred by prior infection and/or vaccination. A version of the test-negative study design (TND), adapted from vaccine effectiveness studies, has been used to assess this relationship. However, the conditions under which such a study identifies the relationship between immune measurements and protection have not been defined.

**Objective:** To evaluate the conditions for TNDs to estimate the relationship between antibody levels or a similar scalar measurement of immunity (hereafter exposure-proximal correlates of protection, COP) and the relative incidence rate of infection given exposure.

**Method:** Individual-based transmission models, linking infection risk linearly and nonlinearly with COP value and accounting for waning immunity post-vaccination and –infection, were used. Simulations were performed of a TND with sampling on predetermined dates. Data from either one or multiple simulation days were analyzed using logistic regression and generalized additive models.

**Result:** A correctly specified logistic regression model provided an unbiased estimate of the effectiveness of specific COP levels (analogous to vaccine effectiveness). Aggregating data across different simulation dates with incidence-density sampling also provided reliable estimates of protection. When, as is generally the case, the functional form relating COP level to protection is unknown, generalized additive models offer a more flexible alternative to traditional logistic regression approaches.

**Conclusion:** A TND can validly estimate the relative effect of an immune COP at the time of exposure on the incidence rate of infection via logistic regression if the functional form of the effect is known and appropriately modeled or unknown a semiparametric approach. Future research should further examine the dynamics of immunity waning and boosting for more reliable inference.

## 1. Introduction

Correlates of protection (COP) are measurable quantities such as binding or neutralizing antibody concentrations that predict the degree of protection against incidence of an infectious disease. These markers provide valuable insights into the immune system’s response to pathogens and vaccines,^1^ which is essential for advancing the understanding of immune mechanisms, as well as facilitating estimates of levels of protection in the population over time and informing the evaluation of new vaccines. For example, hemagglutination-inhibition antibody titers have been identified as a COP for influenza.^2,3^ COP are particularly useful for estimating vaccine effectiveness by linking the magnitude of an immune response to levels of protection, especially in scenarios where direct measures of effectiveness are not available.^4,5^

Recent studies highlight the importance of post-immunization antibody titers as effective COP for COVID-19 vaccines.^6,7^ Notable research efforts have investigated the use of these correlates to forecast absolute risks (AR) and relative risks (RR) of infection in randomized vaccine efficacy trials,^4,6,8–10^ using the antibody concentration measured at a fixed time post-vaccination. More recently, an observational study using a test-negative design (TND) was employed to estimate “exposure-proximal” COP, that is, how the COP level around the time an individual may be exposed to infection affects their risk of becoming infected.^11,12^

The use of the TND for exposure-proximal COP studies builds on a longstanding tradition of using these studies, which compare vaccination histories of those who test positive for a condition (e.g., COVID-19) with those experiencing the same symptoms but testing negative for the condition, to evaluate vaccine effectiveness. If the vaccine provides all-or-nothing protection and there are no unmeasured confounding with respect to infection or test-seeking, such as due to heterogeneous vaccination decisions or characteristics leading to varying susceptibility among participants, the odds ratio (OR) for vaccination among test-positive vs. test-negative participants in a TND is an unbiased estimator of the incidence rate ratio (IRR) in the population, such that one minus the OR estimates vaccine effectiveness (1-IRR). These assumptions are strong,^13^ and methodological work has highlighted that they may be violated in practice.^14–20^

However, to our knowledge, no study has yet evaluated the validity of the TND for monitoring the relationship between a continuous exposure-proximate COP and protection. In contrast with using the TND for vaccine effectiveness, the goal is not to estimate the causal effect, as interventions on the correlate may not be well defined, but rather to estimate the (predictive) relationship between COP-level and infection over time. In this case, our main concern is the potential for selection bias due to the sampling scheme of the TND.

Here, we introduce a novel, simplified simulation model that mimics disease transmission within a community, taking into account vaccination and the waning immunity from previous infections, to explore whether the TND is able to recover the relationship between COP and infection and to identify the correct transformations and statistical models for accurately linking COP to infection. We are specifically interested in situations where assessing vaccine effectiveness in TND may otherwise introduce bias such as when the vaccine exhibits leaky protection. When the relationship between COP, the predictor, and infection risk is linear, we show that a model linking the *ln*(*IRR*) of infection to either the logarithm of one minus the linear predictor of incidence rate (parametric) or a flexible function of that predictor (semiparametric) can effectively recover the correct relationship between that predictor and the incidence rate ratio of infection. When the relationship is unknown, we show semiparametric methods are superior.

## 2. Methods

### 2.1 Simulation model and assumptions

We employed an individual-based transmission model, involving susceptible, exposed, asymptomatically infectious, symptomatically infectious, vaccinated, and recovered individuals, to simulate disease transmission within a community (**Figure 1**), with detailed parameters listed in **Table 1**. Individuals were tracked because each individual had a level of a scalar measure of immunity, denoted as *X*, that varied over time and affected their risk of becoming infected. The model made several key assumptions: The risk of infection decreases linearly as the individual’s COP level increases, calculated as *β* ∗ (*1* − *X*) ∗ (*I* + *A*)/*N*, where *β* is the transmission coefficient, *X* is a rescaled COP level ranging from 0 to 1, *I* is the number of symptomatically infectious individuals, *A* is the number of asymptomatically infectious individuals, and *N* is the total population. One third of the infectious individuals are assumed to be symptomatic and this proportion did not vary over time or across subgroups.^21^ We assume COP provides equal protection against asymptomatic and symptomatic infection. Individuals cannot receive vaccinations if they are symptomatically infectious. All susceptible individuals start with a COP level at 0 units, and a first-time exposure, including recovery after infection or vaccination, will raise the COP level from 0.00 to 0.75 units. With subsequent exposures, this level will boost from current level to 1 unit. These specific values act as simplified indicators to assess the degree of protection conferred against infections following the first^22,23^ and subsequent^24–26^ exposures, respectively. Additionally, all uninfected individuals experience a linear immunity decline at a rate of 0.01 units per day, modeled as a simplified waning mechanism, whereas exposed and infectious individuals will remain their antibody level unchanged until they recover.^27–30^ Data on each individual’s antibody level and infection status is recorded on predetermined simulation dates. In the simulation, 0.5% of the eligible population will be vaccinated every 15 days from day 1 to day 600, totaling 40 rounds. By the end, 20% of the eligible population will receive the vaccine.

**Figure 1.**
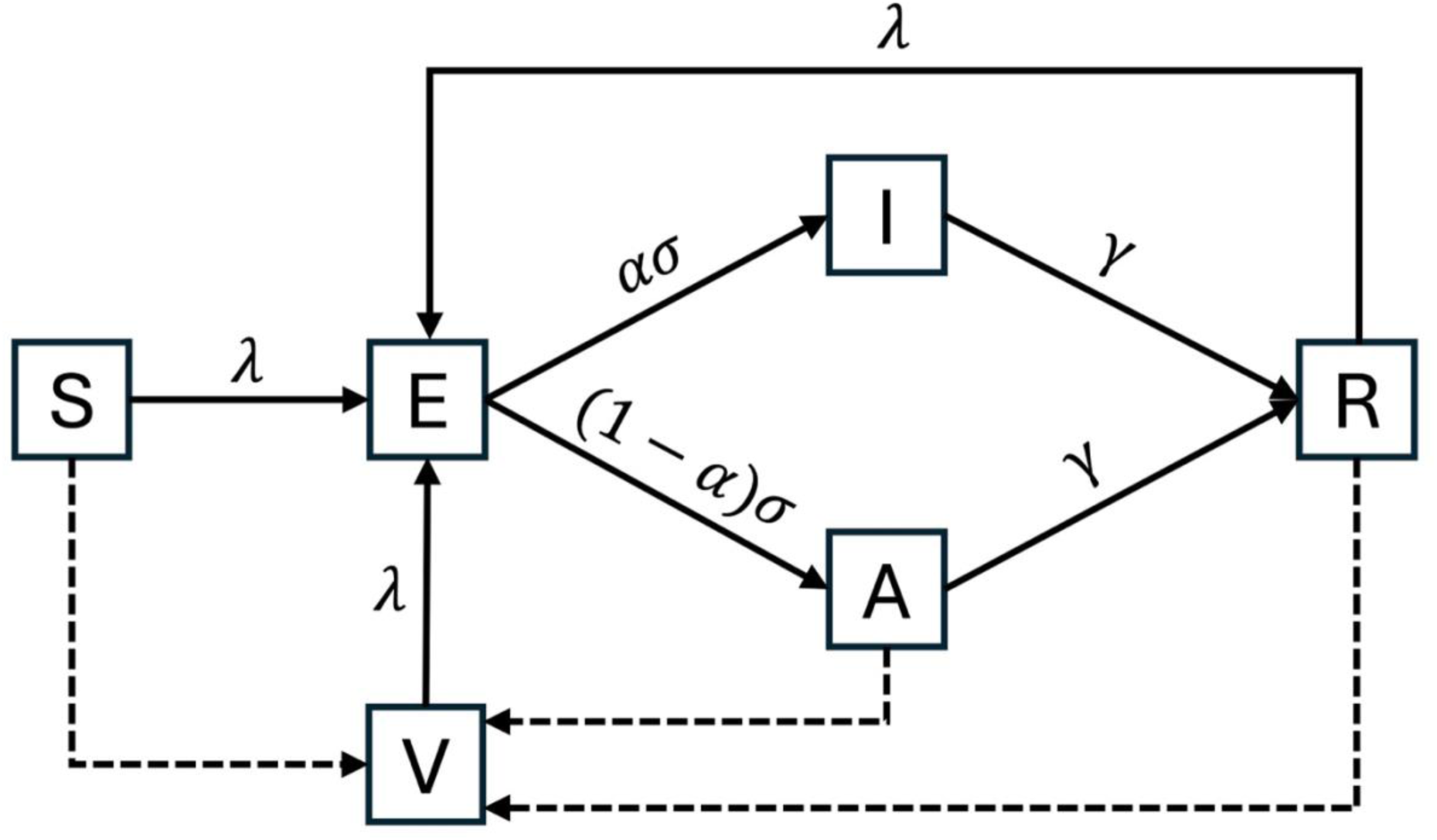
Schematic representation of the model^a^. ^a^ For detailed descriptions of the parameters used, refer to **Table 1**. **State**: S, Susceptible; E, Exposed; I, Symptomatically Infectious; A, Asymptomatically Infectious; R, Recovered; V, Vaccinated. **Arrow**: Solid arrows represent continuous transitions between states; dashed arrows represent the discrete vaccination schedule as detailed in Methods.

**Table 1.** Model parameters.

| Parameter | Description | Value |
| --- | --- | --- |
| N | Population size | 50,000 |
| $t_{\text{latent}}$ | Latent period | 5<sup>31</sup> |
| $t_{\text{infectious}}$ | Infectious period | 7<sup>32</sup> |
| $\alpha$ | Symptomatically infectious proportion | 1/3<sup>21</sup> |
| $\sigma$ | Rate from exposed to infectious | 1 / $t_{\text{latent}}$ |
| $\gamma$ | Rate from infectious to recovered | 1 / $t_{\text{infectious}}$ |
| $R_0$ | Basic reproduction number | 2<sup>33</sup> |
| $\beta$ | Transmission coefficient | $R_0 / t_{\text{infectious}}$ |
| $\lambda$ | Per capita rate of infection | $\beta$ * no. of infectious people |
| $w$ | Antibody level waning rate | 0.01 |
| Initial S | No. of initial susceptible cases | 48,950 |
| Initial E | No. of initial exposed cases | 25 |
| Initial I | No. of initial infectious cases | 25 |
| Initial R | No. of initial recovered cases | 1000 |

### 2.2 Data sampling scheme

In our simulation, we implemented a data sampling scheme modeled after a typical TND study.^34^ Symptomatically infectious individuals were identified as cases, and each case was matched with four controls who were susceptible, recovered, or exposed. This sampling scheme implies selection of controls is independent of COP level on that day. It was assumed that all symptomatically infectious individuals tested positive, and the matched controls tested negative. To increase sample size by aggregating data across multiple days, we used an incidence density sampling approach, matching cases and controls based on the day of sampling.

### 2.3 Statistical analysis

Our analysis considered four regression models: two logistic regression models (with and without transformation of the independent variable, the COP measurement *X*) and two generalized additive models (GAMs, with and without transformation of *X*). COP levels were assessed at the time of sampling, reflecting the exposure-proximate COP — the levels measured when individuals tested positive or negative.

Untransformed models modeled *logit* (*p*) ∼ *X* for logistic regression and *logit* (*p*) ∼ *GAM*(*X*) for the GAM, while transformed models considered *logit* (*p*) ∼ *ln*(*1* − *X*) for the transformed logistic regression and *logit* (*p*) ∼ *GAM*(*ln*(*1* − *X*))for the transformed GAM. The rationale for the transformation was as follows: The transformation applied to these levels was the natural logarithm of 1 – COP level, *ln*(*1* − *x*), which is used to reflect the log of IRR expression in our model setting when comparing the infection incidence rate at a specific COP level to the rate at zero COP level. The derivation is as follows: *ln*(*IRR*) = *ln*{[λ ∗ *I*(*t*) ∗ *U*(*t*) ∗ (*1* − *X*_*X*=*x*_)]/[λ ∗ *I*(*t*) ∗ *U*(*t*) ∗ (*1* − *X*_*X*=_*_0_*)]} = *ln*(*1* − *x*) where λ is the force of infection, *I*(*t*) is the number of infectious individuals at time t, and *U*(*t*) is the number of uninfected individuals at time t. Following the logic of TND vaccine studies, in which the OR provides an unbiased estimate of the IRR of infection among vaccinated versus unvaccinated individuals,^13^ we posited that the log of IRR is linearly related to the log OR comparing individuals with a given level of COP to those who have a COP value of zero and hence the log odds.

Further, the two different infection risk functions—squared (*risk* = *β* ∗ (*1* − *X*^2^) ∗ *I*/*N*) and cubic (*risk* = *β* ∗ (*1* − *X*^3^) ∗ *I*/*N*) transformations of the COP level—were sensitivity analyses used to assess the robustness of the approach to estimating the shape of the relationship between the COP value and incidence when the relationship is more complex and nonlinear.^35,36^ For each of the parametric and semiparametric methods, three models are applied: an untransformed misspecified model, a transformed misspecified model, and a transformed correctly specified model. In both the misspecified and correctly specified transformed models, a natural logarithmic transformation is used. The misspecified transformed models apply linear COP levels, consistent with the primary analysis, while the correctly specified models use squared and cubic COP levels to align with the corresponding infection risk functions.

## 3. Results

The model simulated the spread of infection over 600 days within a community of 50,000 individuals. **Figure 2** shows the temporal dynamics of infectious-and-symptomatic prevalence, COP level proportions, and odds of infection stratified by COP levels. Four complete waves of infection occurred. Notably, declines in the number of individuals with higher antibody levels, due to waning immunity, preceded the emergence of subsequent infection waves. As expected, the simulation shows lower mean and variability in infection odds and lower amplitudes among those with higher COP levels.

**Figure 2.**
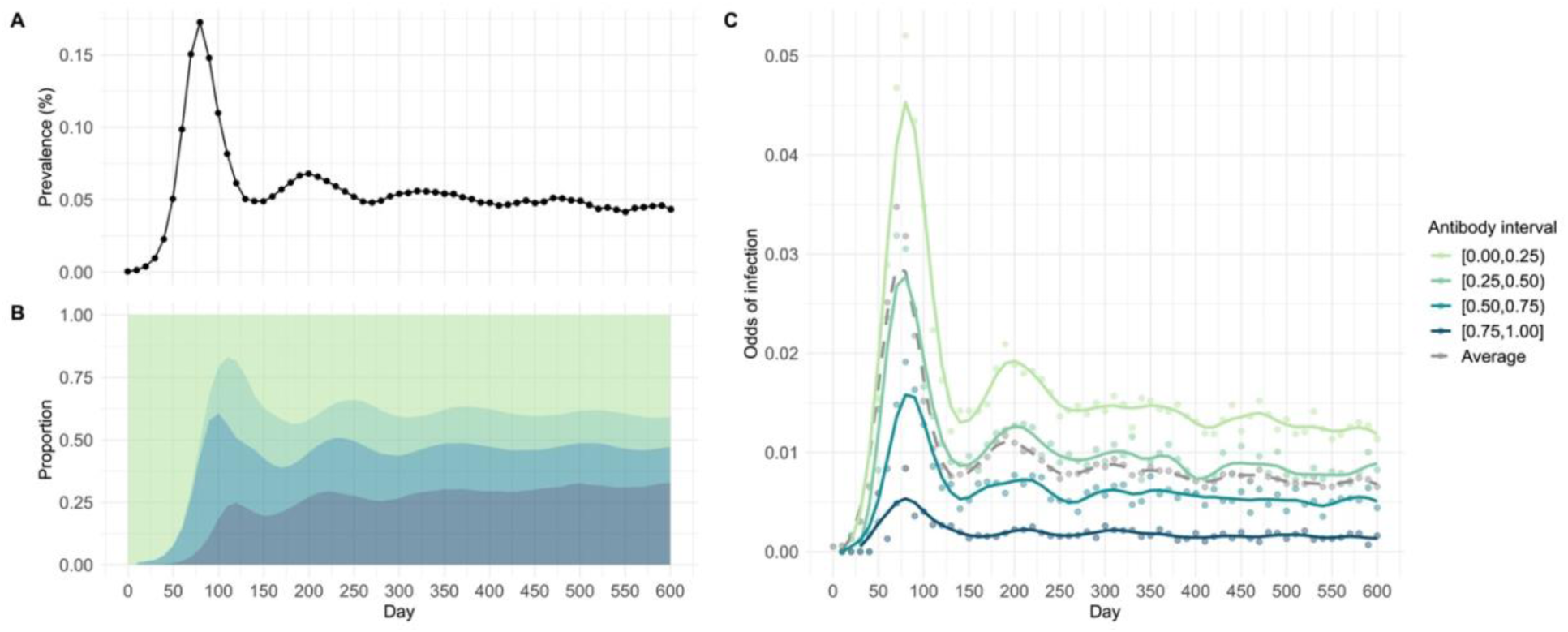
Trends in infectious prevalence, COP proportions, and odds of infection^a^ from simulation days 10 to 600 with an increment of 10. ^a^ The odds of infection curves are fitted using cubic splines with 15 degrees of freedom. **Panel A**: the prevalence of infectious individuals. **Panel B**: the distribution of the population across different antibody intervals. **Panel C**: the odds of incident infection by COP intervals.

TND sampling from the simulation showed that when using data collected from a single day, the transformed logistic model accurately recovered the linear relationship between COP level and IRR. In contrast, the untransformed logistic model failed to do so due to misspecification of the functional form (**Figure 3**). Although both semiparametric GAM models generally captured the relationship, they often deviated from the true pattern, particularly at extreme COP values (**Figure 3A**). **Figure 3B** illustrates the relationships predicted by these models using compiled incidence density data sampled over multiple days, showing that both GAM models produced results more closely aligned with those of the transformed logistic model.

**Figure 3.**
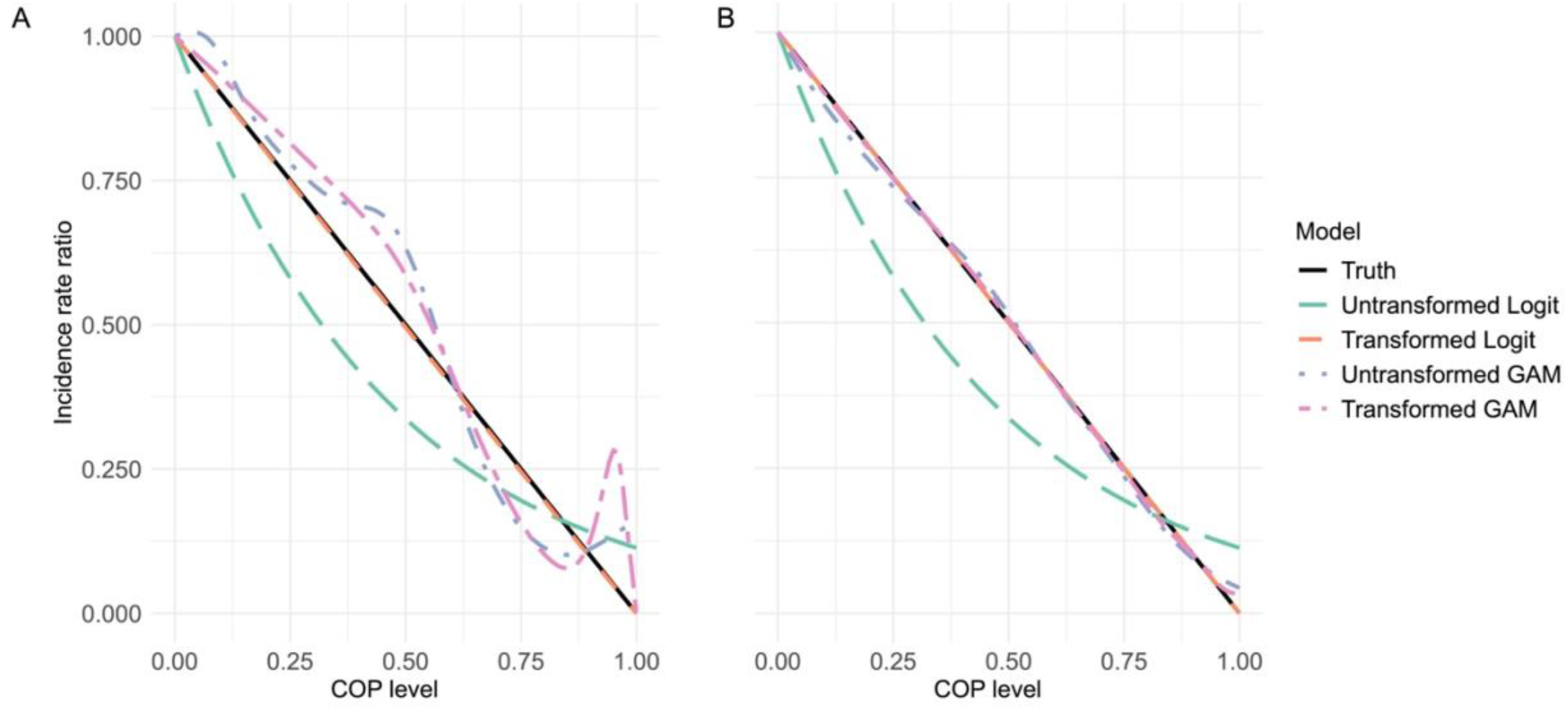
COP level and infection incidence rate ratio sampling on a single simulation day and compiled simulation days. **Panel A** used data collected from simulation day 600, **panel B** used data from simulation days 500-600 with an increment of 10.

The impact of population-level antibody distribution and the aggregation of data from multiple days on the accuracy of regression models in predicting infection IRR were investigated. Besides the transformed logistic regression, both GAMs accurately estimate the IRR when sufficient data are aggregated from multiple days using incidence-density sampling (**Figure 4D**). This accuracy is achieved during periods when the pandemic has stabilized after an extended onset, resulting in a diverse and adequately distributed range of COP levels. In situations with more limited numbers of participants with certain COP levels (**Figure 4A, C**) or insufficient data (**Figure 4A, B**) from TND sampling, semiparametric approaches tend to produce more complex estimated relationships, likely due to overfitting the noise in the data. In contrast, the transformed logistic regression consistently recovers the linear relationship.

**Figure 4.**
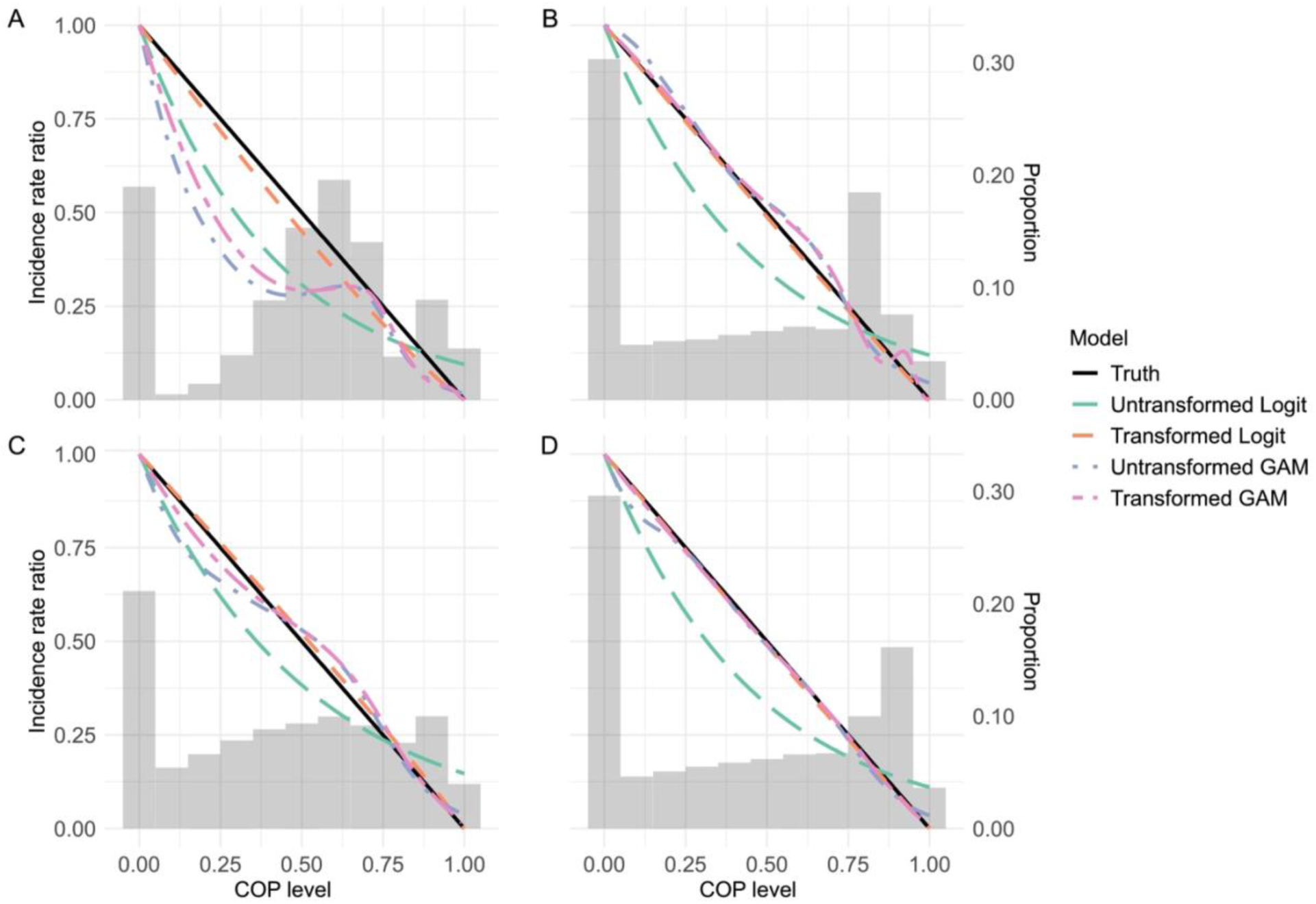
Impact of antibody distribution^a^ and data aggregation on infection incidence rate ratio predictions by regression models. ^a^ Gray areas in the plots indicate the proportions of COP levels within the population. **Panel A** used data collected from simulation day 50, **panel B** used data collected from simulation day 450, **panel C** used data from simulation days 50-150 with an increment of 10, and **panel D** used data from simulation days 400-500 with an increment of 10.

The sensitivity analyses examined the predictability of models on the nonlinear relationships between infection risk and COP levels, including squared and cubic, with sufficient data sampled. The curves generated from the correctly specified transformed logistic regression models are capturing the true relationships, as the primary analysis did, which further shows the robustness of the model selection and transformation function. Although the semiparametric models (whether transformed, correctly specified, or not) did not capture the exact relationship at all levels of COP, they closely approximate the true relationship with only slight fluctuations around the actual lines (**Figure 5**).

**Figure 5.**
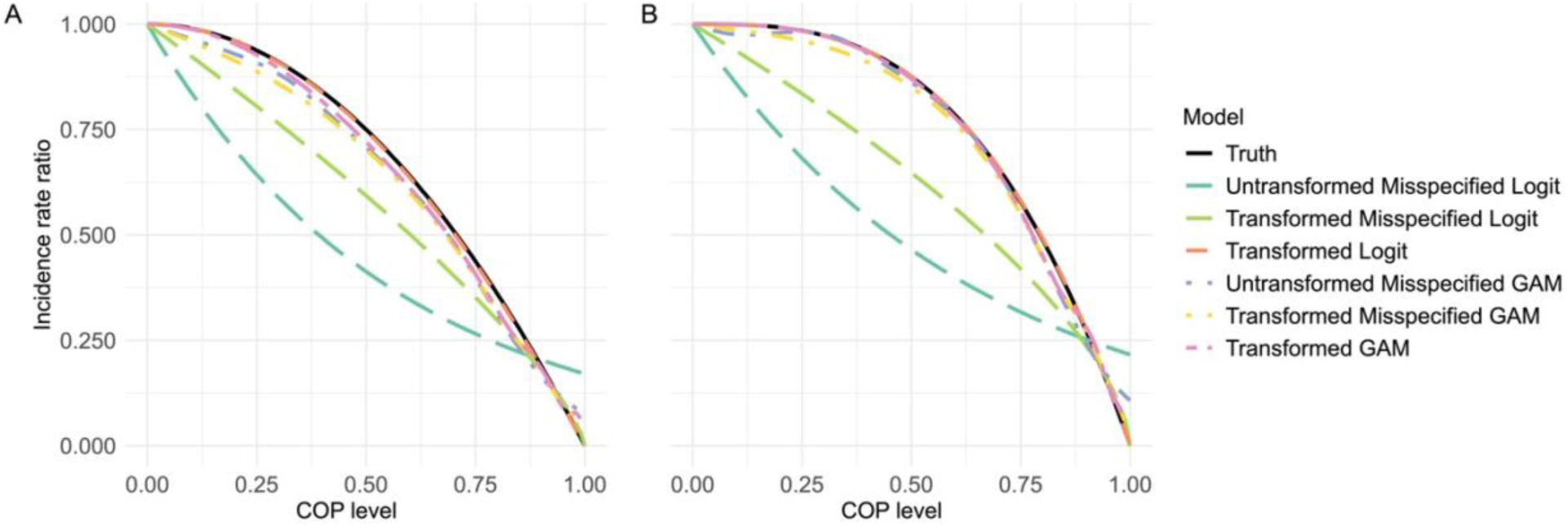
Sensitivity Analysis – COP level and infection incidence rate ratio sampling on compiled simulation days 500 to 600 with an increment of 10. **Panel A** used data from simulation where infection risk correlates with squared COP level, and **panel B** used data from simulation where infection risk correlates with cubic COP level. Formulas used for **Panel A** include: Untransformed Misspecified Logit: *logit* (*p*) ∼ *X* Transformed Misspecified Logit: *logit* (*p*) ∼ *ln*(1 − *X*) Transformed Logit: *logit* (*p*) ∼ *ln*(1 − *X*^2^) Untransformed Misspecified GAM: *logit* (*p*) ∼ *GAM*(*X*) Transformed Misspecified GAM: *logit* (*p*) ∼ *GAM*(1 − *X*) Transformed GAM: *logit* (*p*) ∼ *GAM*(1 − *X*^2^)

## 4. Discussion

Antibody levels have been used in prior studies to predict the infection risk of influenza and COVID-19, and are considered one of the appropriate COP for investigating the relationship between infection and individual immunity. However, apart from one recent paper designed for cohort studies,^11^ there has been little work to define approaches for estimating exposure-proximate COP, that is how an individual’s instantaneously measured level of immunity predicts their susceptibility to infection at that moment. While prospective cohorts have many advantages, case-control designs such as the TND are widely employed due to their comparatively low cost and feasibility. Thus, we sought to understand the conditions under which a TND would validly estimate the relationship between a COP level and the degree of protection offered by that level.

Our study demonstrates that a TND-style approach can identify the shape of a predictive relationship between a correlate of protection measured near the time of exposure and the risk of infection when using a semiparametric model or when using a correctly specified parametric model with appropriate transformation of the value of the correlate. We have phrased this in terms of prediction rather than causal inference because it is difficult to envision even a hypothetical intervention that would set an individual’s COP value at a certain level, and because for many practical purposes, prediction is the question of interest: how well protected is an individual, given a particular COP value? In a causal setting one would also have to consider confounding in which a predictor of COP value had a causal effect on the likelihood of infection. For example, if occupation were predictive of vaccination (and thus COP level) and outcome, or in vaccine campaigns where elderly or immunocompromised individuals are prioritized for early uptake and exhibit lower protection at the same COP value. We have used a model in which such common causes do not exist.

Borrowing from the theory of TNDs, the OR estimates the IRR for a particular covariate value. In the simulation, we define the risk of infection as one minus the immunity level. Therefore, in a logistic regression framework, the right-hand side, *ln*(*IRR*) or *ln*(1 − *COP*), should estimate the left-hand side *ln*(*odds*), and our simulations confirm that using this functional form produces estimates that are indistinguishable from the input to the simulation. Alternatively, using a generalized additive model can approximate this relationship if one does not know the proper functional form for the relationship of the COP value to the IRR.

In TND, estimating the ‘vaccine direct effect’ for leaky vaccines—those that confer only partial protection to all recipients—is problematic and tends to show declining protection over time. The bias arises because vaccinated individuals continue to experience infections at a reduced rate, while unvaccinated individuals may gain immunity through natural infection.^13^ Over time, this dynamic narrows the infection rate gap between vaccinated and unvaccinated groups, causing the OR to trend toward the null. This pattern fails to accurately reflect the vaccine’s true effectiveness. In contrast, this study shows that examining immunity levels as a predictor of infection incidence rate does not suffer from this bias. By analyzing infection rate at varying levels of COP, this approach measures how immunity, regardless of its source, influences infection likelihood. Since it does not rely solely on vaccination status, using COP levels sidesteps the specific biases introduced by leaky vaccine effects in TND. This method, therefore, offers a framework for understanding how incremental COP levels may influence infection incidence rates, while minimizing the impact of vaccine-specific assumptions.

The utilization of simulation models stands out as a major strength of this study, allowing us to emulate real-life pandemic scenarios with a degree of control over experimental variables and pre-assumed infection relative risk functions that are not typically possible in field studies. This approach enabled us to systematically test different infection IRR functions, both linear and nonlinear, and to assess their impact on the relationship between immunity levels and infection risks. Additionally, by employing several logistic regression models on the results obtained from TND, we have enhanced the robustness and applicability of our findings to actual pandemic conditions. Despite its strengths, our study is not without limitations. The infection IRR and risk functions used were intentionally simplified, which might have affected the granularity and generalizability of our findings. Similarly, the model simplifies assumptions about immunity boosting and waning, which may not fully capture the complexities of immunity development and decline in diverse populations. Prior studies suggest that immunity waning could be nonlinear,^35,36^ indicating that the model might overlook important variations. Furthermore, the assumption of homogeneity among agents—considering them to have identical susceptibility and transmission characteristics—may not truly reflect the variability observed in real populations.^37^

Future research should aim to incorporate more realistic infection IRR and risk functions and more sophisticated mechanisms for modeling immunity waning and boosting. Additionally, enhancing the model by calibrating it with real-world data that includes detailed agent characteristics and distributions of COP among circulating variants of concern could significantly improve the model’s accuracy and relevance. Such advancements are important for developing more effective epidemiological models, which, in turn, can inform public health strategies and vaccination programs more accurately.

## 5. Conclusion

Antibody levels are vital in epidemiological research, serving as a key metric for evaluating how these COP are associated with infections under various study designs. These insights are crucial for assessing vaccine efficacy and guiding public health interventions. This study shows that employing logistic regression models with natural logarithm transformations of infection IRR function helps to model the relationship between infection incidence rate and antibody levels more precisely, enabling the visualization of both linear and nonlinear effects. To enhance model accuracy, it is essential to refine infection IRR and risk functions and integrate mechanisms of immunity waning and boosting given vaccination or infection within these models. Calibration with real-world data is crucial to confirm model accuracy and relevance. By transitioning from basic theoretical frameworks to more sophisticated, data-driven models, researchers can more effectively simulate the complex interplay between pathogen exposure, immune response, and population health outcomes, advancing the understanding of immunity dynamics and improving the capacity to predict and manage infectious diseases.

## Data Availability

This is a simulation study with code available at

https://github.com/Ziyuan-Zhang429/COP_IRR.git

